# Human ASXL1 Deficiency Causes Epigenetic Dysfunction, Combined Immunodeficiency and EBV–Associated Hodgkin Lymphoma

**DOI:** 10.1101/2023.12.20.23300096

**Authors:** Maggie P Fu, Mehul Sharma, Sarah M Merrill, Pariya Yousefi, Ryan Tan, Bhavi P Modi, Kate Del Bel, Rebecca J Deyell, Jacob Rozmus, Wingfield Rehmus, Kyla J Hildebrand, Elliot James, Géraldine Blanchard-Rohner, Susan Lin, Kevin E Shopsowitz, Audi Setiadi, Jefferson Terry, Anna F Lee, Britt I Drögemöller, Allison Matthews, Maja Tarailo-Graovac, Laura Sauvé, Hana Mitchell, Julie S Prendiville, Julie L MacIsaac, Kristy Dever, David T S Lin, Mandy Meijer, Colin J D Ross, Simon R M Dobson, Suzanne M Vercauteren, Wyeth W Wasserman, Clara D M van Karnebeek, Margaret L McKinnon, Michael S Kobor, Stuart E Turvey, Catherine M Biggs

**Author notes:** These authors contributed equally (Maggie P Fu, Mehul Sharma). These authors contributed equally (Michael S Kobor, Stuart E Turvey, Catherine M Biggs). **Correspondence to:**Catherine M. Biggs, MD, MSc BC Children’s Hospital, 4480 Oak St, Vancouver, BC, V5Z 4H4, Canada.

## Abstract

Inborn errors of immunity (IEI) are a group of disorders caused by deleterious variants in immune-related genes, including some that function as epigenetic regulators. Additional sex combs-like 1 (ASXL1) is an epigenetic modifier that has not previously been linked to an IEI. Somatic *ASXL1* variants are found in clonal hematopoiesis and hematologic neoplasms, while heterozygous germline variants cause Bohring–Opitz syndrome. We present a new IEI caused by biallelic germline variants in *ASXL1*. The patient had a complex and unusual history of disease progression notable for persistent cutaneous vaccine-strain rubella granulomas initially manifesting in early childhood, chronic macrocytosis and mild bone marrow cellular hypoplasia, and Epstein Barr virus– associated Hodgkin lymphoma in adolescence. Detailed immunophenotyping revealed progressive loss of B-cells, hypogammaglobinemia, and T-cell lymphopenia with severe skewing toward a memory phenotype and elevated expression of T-cell exhaustion and senescence markers. Molecular investigations confirmed ASXL1 protein deficiency in the patient’s T-cells and fibroblasts. The T-cells exhibited marked loss of DNA methylation, increased epigenetic aging, and CD8 T-cell dysfunction. These aberrations were ameliorated by lentivirus-mediated transduction with wild-type *ASXL1*, confirming the pathogenicity of *ASXL1* variants. This study defines a novel human IEI caused by ASXL1 deficiency, a diagnosis that should be considered in individuals with chronic viral infections, virus-associated hematologic malignancies, and combined immunodeficiency. Furthermore, our findings provide fresh insights into the mechanisms underlying the roles of human ASXL1 in T-cell function as well as in the development and maintenance of lymphomas.

## INTRODUCTION

Monogenic inborn errors of immunity (IEIs) are a group of more than 480 rare diseases caused by deleterious germline variants in immune-related genes.^1^ Some of these IEI disease-causing variants occur in genes encoding epigenetic regulators that modify steady-state gene expression and cellular phenotype, such as *DNMT3A* and *TET2*.^2,3^ These alterations in epigenetic regulation disrupt hematopoiesis and cellular differentiation pathways that are important for immune function,^4,5^ giving rise to inflammation, lymphoproliferation, and increased susceptibility to infections and cancer.^6,7^

Hematologic malignancies account for the majority of cancers in IEIs and are mainly of lymphoid origin, however, myeloid leukemias and myelodysplastic syndromes (MDS) also occur.^6^ Impaired cell-mediated immunosurveillance, chronic viral infections, and inflammation contribute to the increased risk and early onset of cancers observed in IEIs.^8,9^ Furthermore, the same mechanisms underlying immunodeficiency in certain IEIs may also promote oncogenesis in a cell-intrinsic manner, such as DNA repair or epigenetic defects, causing genomic instability.^9^ Somatic variants in IEI-associated genes are also observed in hematologic malignancies, further supporting their oncogenic role.^10^ Indeed, acquired variants in the epigenetic regulatory genes *DNMT3A*, *TET2*, and *ASXL1* (DTA mutations) are common in age-related clonal hematopoiesis and myeloid neoplasms.^11,12,13,14^ *ASXL1* (MIM #612990) is the least characterized of the three DTA genes, and unlike *DNMT3A* and *TET2* has not previously been linked with immunodeficiency in humans.

ASXL1 comprises part of the Polycomb repressive deubiquitinase (PR-DUB) complex,^15^ which along with Polycomb repressor complexes 1 and 2 (PRC1/2) represses target genes by modifying chromatin. In embryonic development, establishment of histone H2A lysine 119 monoubiquitination (H2AK119ub1) by PRC1 is crucial for the recruitment of PRC2 and corresponding transcriptional repression, and plays a key role in establishing chromatin bivalency whereby histone modifications associated with transcriptional activation or repression co-occur.^16,17^ The PR-DUB complex removes H2AK119ub1 at later developmental stages to maintain transcriptional _regulation.16,17,18,19_

In keeping with the critical role of PR-DUB in development, *de novo* heterozygous loss-of-function (LoF) *ASXL1* variants cause the neurodevelopmental disorder Bohring–Opitz Syndrome (BOS), characterized by severe developmental delay, microcephaly, failure to thrive, and distinct facial features.^20^ While *Asxl1*-null mice recapitulate BOS phenotypes, heterozygous germline deletion and hematopoietic-specific *Asxl1* deletion lead to MDS-like features.^21,22^ Mechanistic studies of BOS and ASXL1-associated hematologic malignancies implicate ASXL1 in differential DNA methylation patterns and altered epigenetic age.^23,24,25,26^ Further, conditional knock-in of an *Asxl1* variant associated with clonal hematopoiesis revealed a phenotype of impaired T-cell development, function and exhaustion that drove tumor progression in mice.^27,28,29^

Here, we report the first patient with an IEI due to biallelic germline variants in *ASXL1* causing ASXL1 deficiency presenting with combined immunodeficiency complicated by vaccine-strain rubella virus granulomas and Epstein–Barr virus (EBV)-associated Hodgkin lymphoma. We characterize the patient’s clinical phenotype and treatment course, and provide a mechanistic study of the epigenetic and immunological impact of autosomal recessive ASXL1 deficiency. We anticipate that this discovery will empower the diagnosis and management of additional patients with ASXL1 deficiency.

## METHODS

### Participants and consent

Peripheral blood samples were collected from the index patient and patient’s parents and sibling, as well as three unrelated healthy individuals at the BC Children’s Hospital Research Institute. Similarly, skin biopsies were collected from the patient and six healthy individuals. All participants from whom we collected biological samples and/or their parents/guardians provided written informed consent for sample collection, sequencing, data analysis, and the publication of findings and photographs. Data from additional samples were obtained from publicly available databases, as outlined below. The research protocols were approved by the University of British Columbia Clinical Research Ethics Board (H15-00641).

### Identification of candidate variant

Genomic DNA was isolated from peripheral blood using standard procedures and exome/genome sequencing was performed on the Illumina HiSeq platform (Macrogen, Seoul, South Korea). Sequencing data were analyzed using an updated version of our in-house, open-source, semi-automated bioinformatics pipeline as described previously.^30,31^ Trio whole-exome sequencing (trio-WES) was performed on genomic DNA from the patient and both parents. Compound heterozygous *ASXL1* variants were considered the top candidates of interest. Additional information on candidate gene selection is available in the **Supplementary Methods**. Whole-genome sequencing (WGS) was then performed to scan for potential somatic mosaic variants and to exclude variants in noncoding regions. Compound heterozygous *ASXL1* variants still emerged as the variants of highest significance and were further verified by sequencing (**Supplementary Methods**).

### Generation and maintenance of patient’s primary T-cells and fibroblasts

For generation of T-cell blasts, T-cells were isolated from peripheral blood mononuclear cells (PBMCs) (EasySep™ Human T-Cell Isolation Kit, Cat # 17951; Stemcell, Vancouver, BC, Canada) and activated for 12 days (**Supplementary Methods**).

To generate fibroblasts, a skin biopsy was obtained, manually cut into smaller pieces, and placed in separate wells of a 6-well plate in complete DMEM (GE Healthcare, Chicago, IL, USA) until formation of a confluent fibroblast monolayer (**Supplementary Methods**).

### Stable WT ASXL1 expression in expanded patient-derived T-cells

Stable wild-type (WT) ASXL1 expression was achieved by infection of expanded patient-derived T-cells with lentiviral particles as described previously,^32,33^ and expanded cells were used for downstream DNA methylation and intracellular flow cytometric analyses (**Supplementary Methods**).

### DNA methylation array

Blood and buccal swabs were collected from the index patient, and genomic DNA was extracted from whole blood (WB), PBMCs, buccal cells, and cultured T-cells. Aliquots of 160 ng of extracted DNA (750 ng) subjected to bisulfite conversion (EZ-96 DNA Methylation kit; Zymo Research, Irvine, CA, USA) were assayed. DNA extracted from primary tissues and T-cells was assayed using Infinium HumanMethylationEPIC BeadChip (EPIC) and HumanMethylationEPICv2 BeadChip arrays (Illumina, San Diego, CA, USA), respectively. Two patient PBMC samples were prepared and assayed as technical replicates on the EPIC arrays. (**Supplementary Methods**).

### Sample selection for pediatric population reference set

To establish a pediatric population reference set, publicly available DNAm microarray data on blood and buccal samples were downloaded from the Gene Expression Omnibus (GEO) database, including both EPIC arrays and their predecessors, the Illumina Infinium HumanMethylation450 BeadChip (450k) arrays. We filtered for healthy control (HC) samples with the cohorts and sample selection outlined in the **Supplementary Methods**. The cohorts and demographics are described in **Supplementary Table 1-3**. Details of normalization, quality control, and imputation of the HC and patient samples are also presented in the **Supplementary Methods**.

### Epigenetic age analysis

The index patient, family control, and unrelated HC blood, buccal, and T-cell epigenetic age were estimated with the Horvath pan-tissue clock,^34^ Hannum clock,^35^ and Horvath skin and blood clock^36^ using the methylclock R package (**Supplementary Methods**).

### Differential DNA methylation analysis

Cell-type proportion prediction, subsequent accounting for intersample cellular heterogeneity, and adjusting for other sources of batch effects were performed prior to differential methylation analysis as presented in the **Supplementary Methods**. For differential DNA methylation analysis, linear regression were performed with `limma`.^37^ For each primary tissue, the regression model accounted for sex and age, whereas for cultured T-cells we additionally accounted for batch and CD8 T-cell proportion measured by flow cytometry. Details of the regression model are presented in the **Supplementary Methods**. Multiple test correction was performed with the Benjamini-Hochberg procedure^38^. Skewness of the differential DNAm pattern was assessed with skewness test for normality, implemented with *skewness.norm.test* function in the `normtest` package.^39^

### Enrichment analysis

Overrepresentation analyses were performed to identify enriched ChromHMM chromatin states with the *gometh* function in the missMethyl package (**Supplementary Methods**).

### Intracellular flow cytometry in PBMCs, expanded T-cells, and primary fibroblasts

Intracellular flow cytometry was performed to examine the expression of ASXL1, H2AK119ub1, and IL-2-induced phosopho-STAT5 (pSTAT5) in expanded patient-derived primary T-cells; pSTAT5 in PBMCs; and ASXL1 in primary fibroblasts (**Supplementary Methods**).

### Clinical-grade flow cytometry

Immunophenotyping of clinical samples was performed using WB at an accredited clinical flow cytometry laboratory following established standard protocols (**Supplementary Methods**).

### Histology

Formalin-fixed, paraffin-embedded lymph node tissue was cut into sections 3 µm thick for routine hematoxylin and eosin (H&E) staining or 5 µm thick for immunohistochemistry. Bone marrow trephine biopsies were processed in B-plus solution followed by decalcification and formalin fixation. Paraffin embedded marrow tissue was then cut into sections 3 µm thick for routine hematoxylin and eosin (H&E) staining or 4 µm thick for immunohistochemistry.

### Data availability

The data is available upon request by emailing the corresponding author.

## RESULTS

### Case description

The index patient is currently an adolescent female (10 to 19 years of age) born to nonconsanguineous parents. One parent has a history of rheumatoid arthritis, while the other parent and sibling have no notable clinical history. The index patient was born full-term and received all routine immunizations, including live attenuated measles, mumps, rubella (MMR) vaccine (**Fig. 1A**). During early childhood (0 to 5 years of age), she developed chronic ulcerated skin granulomas on the right arm and left leg. She was hospitalized twice for pneumonia between 0 to 10 years of age, with the latter in the context of primary EBV infection. Laboratory tests over time showed chronic macrocytosis, T-cell lymphopenia, progressive loss of B-cells, and hypogammaglobulinemia (**Fig. 1B, Supp. Fig. 1B**).

**Figure 1:**
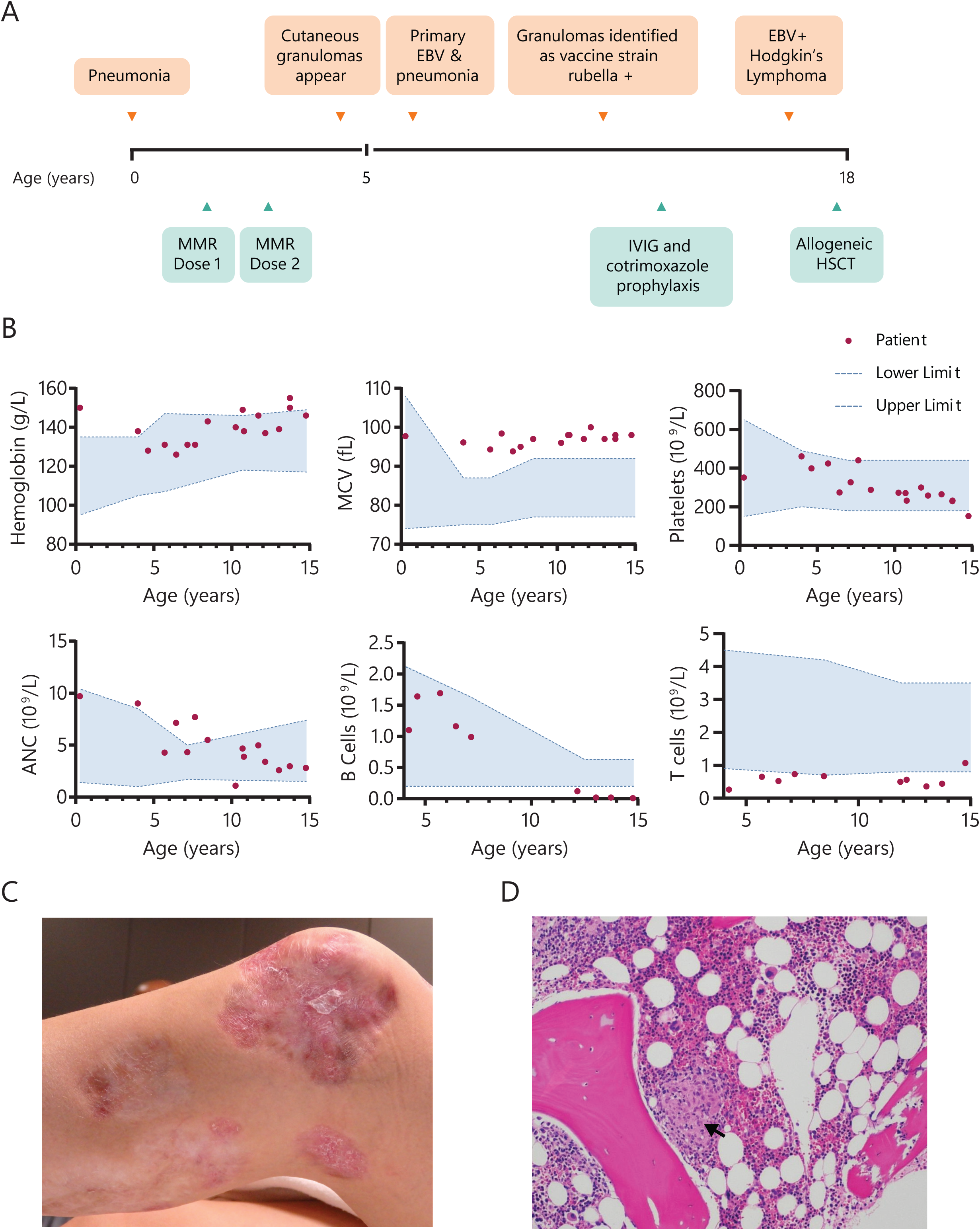
Clinical phenotype of a patient with biallelic *ASXL1* variants. A) Clinical timeline depicting the patient’s infections requiring hospitalization in early childhood (pneumonia and primary EBV infection with bacterial pneumonia), timing of MMR vaccines, onset of cutaneous granulomas and EBV^+^ Hodgkin lymphoma, and definitive treatment with HSCT. B) Timeline of hematologic parameters, demonstrating normal/elevated hemoglobin, declining platelet and absolute neutrophil counts, chronic macrocytosis, low/normal T-cell counts, and progressive loss of B-cells. C) Image of healing cutaneous granuloma on left knee taken in early adolescence (10-15 years of age). D) Bone marrow biopsy showing mildly hypocellular marrow and presence of a granuloma (arrowhead). ANC, absolute neutrophil count; Hb, hemoglobin; HSCT, hematopoietic stem cell transplantation; MCV, mean corpuscular volume; Plt, platelets.

Despite prolonged systemic courses of antimicrobials, topical corticosteroids and wound care, the skin granulomas persisted for more than a decade, with new lesions developing in the left inguinal region. Repeated biopsies showed necrotizing granulomatous inflammation with no organisms identified on special stains. (**Fig. 1C, Supp. Fig. 1C**). Serial bone marrow biopsies every 1–2 years showed mild hypocellularity, megaloblastoid changes, and eventually the presence of non-necrotizing granulomas (**Fig. 1D**). Infectious studies revealed intermittent EBV viremia (ranging from 0 to 1000s copies/mL), absent antibody titers to measles, mumps, varicella, and hepatitis B, but high titers to rubella. A biopsy from one of the cutaneous granulomas was tested for vaccine-strain rubella, RA 27/3, and was found to be positive by polymerase chain reaction.

In early adolescence (10 to 15 years of age), the patient was diagnosed with EBV-associated stage IVA classical Hodgkin lymphoma, mixed cellularity subtype with prominent granulomatous inflammation (**Supp Fig 1D and Supplementary Clinical Data)**. She underwent four cycles of chemotherapy, at which time a repeat Positron Emission Topography (PET)-CT showed progressive disease, and she was transitioned to nivolumab, anti-programmed cell death protein 1 (PD1) antibody, immunotherapy.

This was complicated by the development of autoimmune thyroiditis treated with thyroxine. Six weeks following discontinuation of nivolumab, conditioning was begun for a 9/10 matched unrelated donor hematopoietic stem cell transplant (HSCT) with cyclophosphamide, fludarabine and 200cGy total body irradiation. She received mycophenolate mofetil and tacrolimus prophylaxis for graft versus host disease (GVHD) and developed grade 1 GVHD of upper gut and skin which was treated with corticosteroids. She was discharged on day +34 with full donor chimerism, and remains well now 6 months posttransplant with healing of her chronic skin lesions.

### Identification and characterization of novel germline compound heterozygous *ASXL1* variants in the index patient

Given the unique clinical presentation and the severity of the clinical phenotype, trio-WES and subsequent WGS were performed. Sequencing results identified compound heterozygous variants in *ASXL1*, a maternally inherited variant in exon 5 (Var1: NM_015338.6:c.332G>A;NP_056153.2:p.Cys111Tyr) and a paternally inherited variant in exon 13 (Var 2: NM_015338.6:c.3710C>T;NP_056153.2:p.Ser1237Phe), as candidates. Sequencing confirmed that the biallelic *ASXL1* variants segregated with the presence of disease in the family (**Fig. 2A,B**). These are missense variants predicted to be deleterious, and were considered to be strong candidates for further functional evaluation based on their changes in amino acid polarities, high combined annotation-dependent depletion (CADD) scores (Var1: 26.2; Var 2: 23.0), and their low allelic frequency in the Genome Aggregation Database (gnomAD) v3 (Var1: 8.12e−06; Var2: 1.22e−05, with no homozygotes for either variant; **Supp. Fig. 2A**). Both variants fell outside the mutation hotspot region associated with typical truncating BOS variants in *ASXL1* exon 13 (**Fig. 2C**).

**Figure 2:**
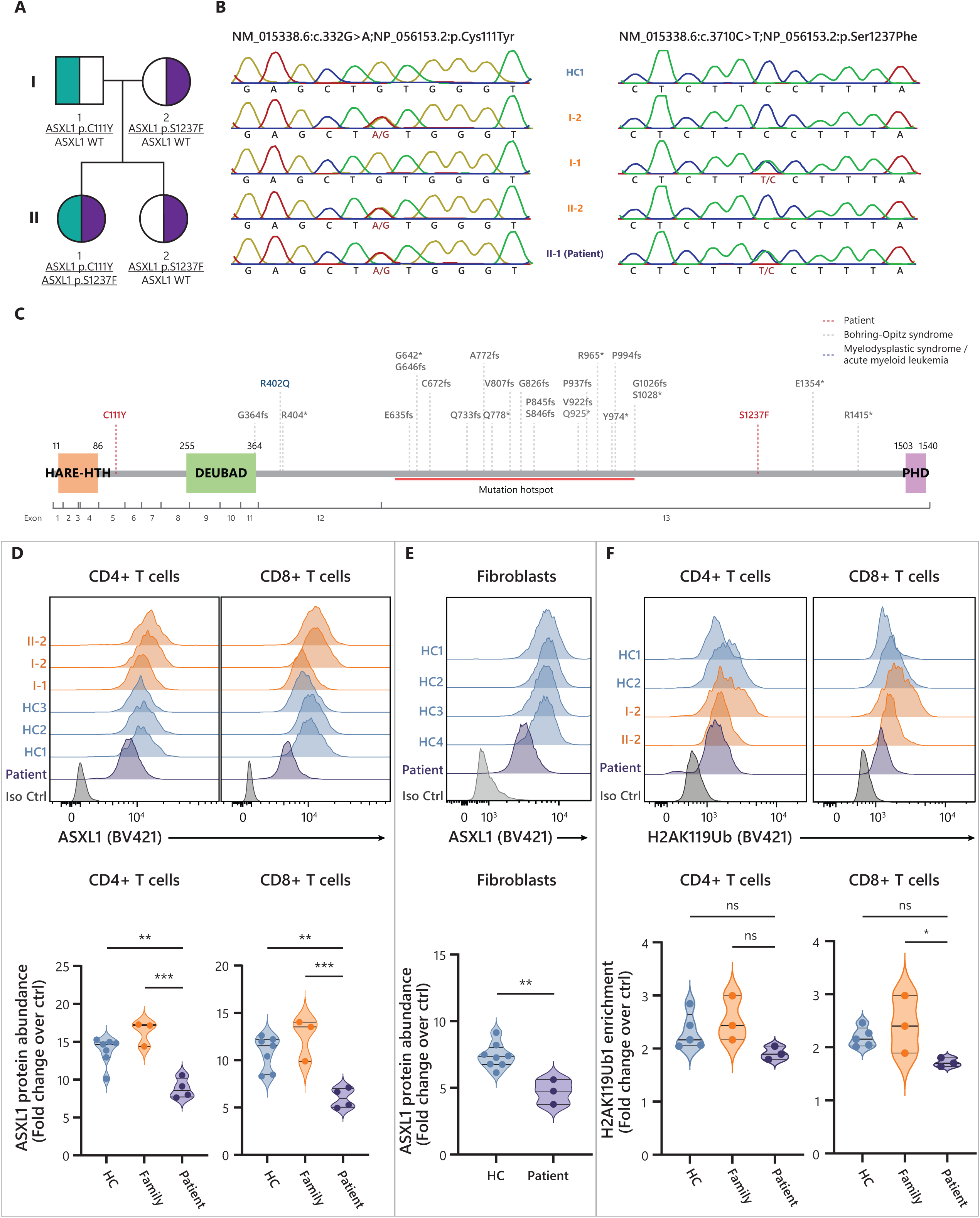
Characterization of germline biallelic *ASXL1* variants identified in the index patient. A) Family pedigree of *ASXL1* variant inheritance pattern. B) Index patient and her family’s *ASXL1* genotype as verified by sequencing. C) Schematic of major functional domains in ASXL1 and the positions of patient and other reported germline variants. Patient’s *ASXL1* variants (C111Y and S1237F, labeled red) are located outside the mutational hotspot and major functional domains. A variant linked to non-Hodgkin lymphoma (R402Q) is labeled blue, and those linked to BOS are labeled gray. D) Histograms of patient and control ASXL1 protein abundance in expanded T-cells (CD4^+^ and CD8^+^) based on flow cytometry, and their respective quantification as measured over four separate experiments. E) Histogram of patient and HC ASXL1 protein abundance in cultured fibroblasts using flow cytometry, and their respective quantification as measured over three separate experiments. F) Histogram of patient and HC H2AK119 ubiquitin levels in expanded CD4^+^ and CD8^+^ T-cells based on flow cytometry, and their respective quantification as measured over three separate experiments. Results from one representative experiment are shown in each of D, E, and F. \**P* < .05, \*\**P* < .01, \*\*\**P* < .001, one-way analysis of variance and Dunnett’s multiple comparison test.

Molecular consequences of the patient’s *ASXL1* variants were investigated next. Expanded patient primary T-cells (both CD4 and CD8) expressed approximately half the amount of total ASXL1 protein when compared to both healthy controls (HCs) and family members (p<0.05, **Fig 2D**). ASXL1 deficiency was also confirmed in patient cultured fibroblasts obtained from a skin biopsy (**Fig. 2E**). Given the well-described role of ASXL1 in H2AK119 deubiquitination,^40^ we quantified H2AK119ub1 in expanded T-cells but did not find notable differences in bulk enrichment levels between the patient and controls (**Fig. 2F**). While surprising, recent work has shown that BOS-causing germline *ASXL1* variants do not alter specific histone modifications, including H2AK119ub1, but rather cause DNAm dysregulation.^23^ Therefore, we next assessed the global DNAm signatures in the patient.

### Differential DNAm in PRC-regulated genomic elements and advanced epigenetic aging in the patient’s lymphocytes

We conducted epigenome-wide association studies (EWAS) to compare the DNAm patterns of primary patient samples to age-matched HCs compiled from publicly available data sets in three tissue types: buccal swabs (*n* = 725), whole blood (WB) (*n* = 452), and isolated PBMCs (*n* = 285). We observed wide-spread differential DNAm between the patient and HCs in WB and PBMCs (**Fig. 3A**) but not in buccal swabs (**Supp. Fig. 3A**). There were 50 885 differentially methylated sites (DMs) in WB, 18 581 DMs in PBMCs, but only 38 DMs in buccal swabs that passed the threshold of adjusted *P*-value (*P*_adj_) ≤ .05, 15.7% of which (9438 DMs) overlapped between WB and PBMCs (**Fig. 3B**). Emphasizing that this patient does not have BOS, we found that BOS-associated DMs were not enriched in our analysis (**Supp.** Fig. 3B). Pathway enrichment analysis with 18-state ChromHMM chromatin state annotation showed that DMs in both WB and PBMCs were significantly enriched in regulatory elements, such as enhancers, bivalent transcription start sites, and repressed Polycomb regions, as well as some quiescent gene and heterochromatin regions in PBMCs (**Fig. 3C**).

**Figure 3:**
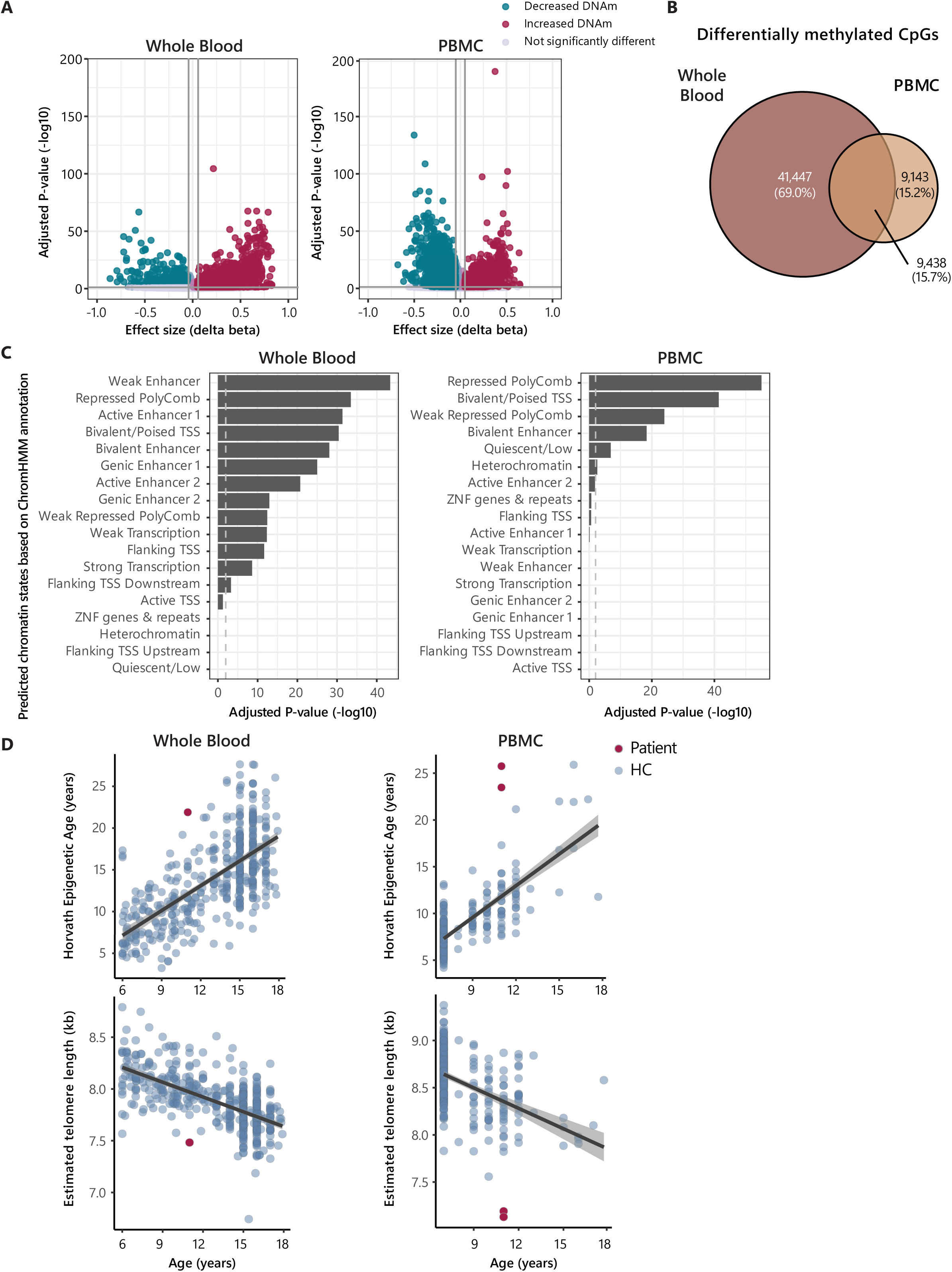
Patient and HC DNAm profiles and EA. A) Volcano plot of differential DNAm patterns detected in WB and PBMCs with linear regression. Δβ represents the effect size of the patient status variable after accounting for age at sample collection and sex. Magenta points indicate DMs with increased DNAm in the patient, whereas the turquoise points represent those with decreased DNAm. The vertical dark gray line represents an effect size threshold of 0.1, and the horizontal line corresponds to the statistical threshold of *P*_adj_ = .05. B) Venn diagram showing the overlap of DMs with *P*_adj_ < .05 in WB and PBMCs. C) Enriched chromatin states based on the DMs in WB and PBMCs using the 18-state ChromHMM model. Overrepresentation analysis was used to extrapolate the significance of the enrichment. The gray dashed line corresponds to *P*_adj_ = .05. D) Horvath pan-tissue EA was calculated in WB and PBMCs with the methylclock package. HC samples are indicated in light blue, while patient samples are shown in red. The black line is the linear model best fit line. The gray area corresponds to the 95% confidence interval.

To gain further information about the patient’s DNA methylome, we assessed the patient’s tissue-specific EA. In analysis of buccal swabs, the patient was estimated to have a pediatric buccal epigenetic (PedBE) clock age of 10 to 15 years, matching her actual age of 10 to 15 years at sample collection (**Supp. Fig. 3C**). In contrast, the Horvath pan-tissue age of blood samples was higher compared to her chronological age (**Fig. 3D**). Similarly, the predicted telomere length based on DNAm data was also lower in blood (**Fig. 3D**), and the presence of short telomeres in lymphocytes and granulocytes was confirmed by direct measurement (**Supp. Fig. 3D**). Overall, the ASXL1-deficient patient had increased EA and short telomeres compared to HC, which was especially prominent in lymphocyte subsets.

### Wide-spread DNAm loss and advanced epigenetic aging in patient’s T-cells were rescued by WT *ASXL1* transduction

We transduced expanded patient-derived T-cells with WT *ASXL1* to delineate the relations between the patient’s variants and the alterations in epigenetic regulation in lymphocytes. Whereas empty vector (EV)-transduced patient cells (patient condition) maintained a lower ASXL1 protein level compared to the EV-transduced HC cells (HC condition), WT *ASXL1*-transduced patient cells (rescued condition) showed increased ASXL1 protein comparable to the HC condition (**Fig. 4A**).

**Figure 4:**
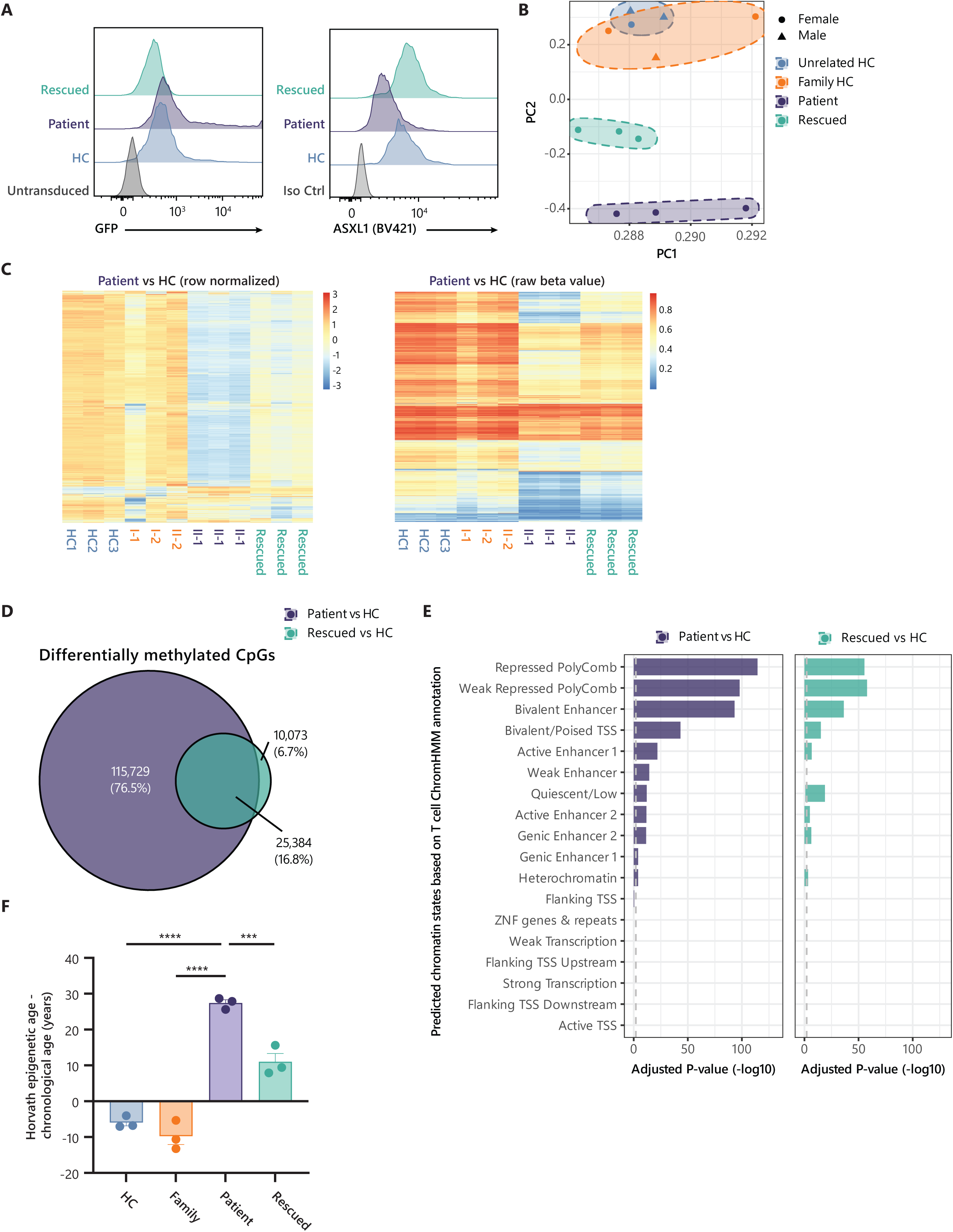
DNA methylation disturbance in expanded patient-derived T-cells and molecular rescue by WT *ASXL1* transduction. A) GFP signal and ASXL1 protein abundance in expanded T-cells transduced with either EV or WT *ASXL1*, as detected by flow cytometry. The vectors contained a GFP tag, so the transduction efficiency was represented by the GFP signal. The HC condition consisted of expanded HC T-cells with EV transduction, while the patient and rescue conditions consisted of expanded patient-derived T-cells transduced with EV or WT *ASXL1*, respectively. B) Biplot of PC1 and PC2 generated by PCA of DNAm measured at all loci on the methylation microarray. Unrelated HC samples are labeled blue, family HCs are labeled orange, patient samples are labeled purple, and rescued samples are labeled green. C) Heat map of DMs in linear regression analysis comparing patient with unrelated and family HC samples, with the DMs in rows and samples in columns. The heat map on the left is row normalized, and that on the right shows the DNAm level (beta value). D) Venn diagram showing the DMs in the *patient vs. HC* (purple) and *rescued vs. HC* (green) comparisons. The DMs were detected with thresholds of *P*_adj_ < .2 and delta-beta > 0.1. E) Enriched chromatin states based on the DMs in the *patient vs. HC* (purple) and *rescued vs. HC* (green) comparisons using the 18-state ChromHMM model. Overrepresentation analysis was used to extrapolate the significance of the enrichment. The gray dashed line corresponds to *P*_adj_ = .05. F) Bar graph showing the differences in Horvath pantissue epigenetic age and chronological age of the expanded T-cells. \**P* < .05, \*\**P* < .01, \*\*\**P* < .001, one-way analysis of variance with Dunnett’s multiple comparison test. GFP, green fluorescent protein; Iso ctrl, isotype control

Principal component analysis (PCA) of the T-cells’ DNAm showed that the patient clustered away from the HC (including family member) cells along PC2, with the rescued cells in the middle (**Fig. 4B**). Analysis of the *patient vs. HC* comparison yielded 141 113 differentially methylated sites (DMs) (*P*_adj_ < .2 and delta-beta > 0.1), of which 129,104 (91.5%) showed decreased DNAm. The pattern observed in the PCA plot was replicated in the DMs—patient samples were notable for their decreased DNAm, while the rescued condition showed an intermediate DNAm level (**Fig. 4C**). Furthermore, loss of DNAm in the patient condition occurred mainly in loci that were highly methylated in HC samples; of the DMs with decreased DNAm, 80.1% were > 50% methylated in the HCs (skewness test *P* < 2.2e−16, **Fig. 4C**). In the *rescued vs. HC* condition, 35 457 DMs were identified, and 90 354 of the *patient vs. HC* DMs were not significantly different in this comparison, indicating that the majority of the DMs in the patient’s cells were rescued by WT *ASXL1* transduction (**Fig. 4D**). Similar to our findings in blood, the genomic locations of DMs were highly enriched for repressed Polycomb and bivalent chromatin states, with the rescued cells showing higher *P*-values than that of patient cells (**Fig. 4D,E**).

We also investigated epigenetic aging in this context. For the controls, the Horvath pan-tissue clock slightly underpredicted chronological age. In contrast, the clock assigned the patient an epigenetic age on average 27.4 years older than her chronological age, and WT *ASXL1* lowered her epigenetic age (**Fig. 4F**). Specifically, the difference between epigenetic and chronological age was significantly higher in the patient than family and unrelated HCs, and this difference was significantly reduced by WT *ASXL1* (*P* < .0005) (**Fig. 4F**). Other epigenetic clocks corroborated the trend (**Supp. Fig. 4A,B**). Overall, WT *ASXL1* transduction rescued the DNAm differences and attenuated the advanced epigenetic aging driven by the patient’s *ASXL1* variants.

### *ASXL1* variants caused impaired CD4 and CD8 T-cell maintenance and activation in patient primary cells

Given the methylation impairments in patient T-cells, we conducted detailed immunophenotyping of peripheral blood prior to the patient’s lymphoma diagnosis. Naïve CD4 and CD8 T-cell subsets and recent thymic emigrants were markedly reduced, suggesting poor thymic output (**Fig. 5A**, **Table 1**). The patient showed expansion of PD1^+^ CD4 and CD8, and CD57^+^ CD8 T-cell populations (**Fig. 5A**), indicating T-cell exhaustion and senescence. CD8 T-cells had a restricted T-cell receptor repertoire (**Table 1**). We then assessed pSTAT5 signaling and reactive oxygen species (ROS) production, as alterations in these pathways were reported in T-cells containing an *ASXL1* variant associated with clonal hematopoiesis of indeterminate potential.^29^ We observed significantly increased pSTAT5 abundance after low-dose IL-2 (10U/mL) treatment in naïve CD8 T-cells (**Fig 5B**) but no difference in the memory T-cell compartment (**Supp Fig 5A-B**). Reduced pSTAT5 production was observed in expanded patient CD8 T-cells after IL-2 treatment, which was rescued by stable WT *ASXL1* expression, indicating a causative role for ASXL1 in regulating STAT5-mediated CD8 T-cell homeostasis (**Fig 5C**). Finally, high ROS production was observed in expanded patient CD4 T-cells, which was rescued upon transduction of WT *ASXL1* (**Supp Fig 5C**), suggesting alternative modes of *ASXL1* variant-mediated impairments of CD4 and CD8 T-cells.

**Figure 5:**
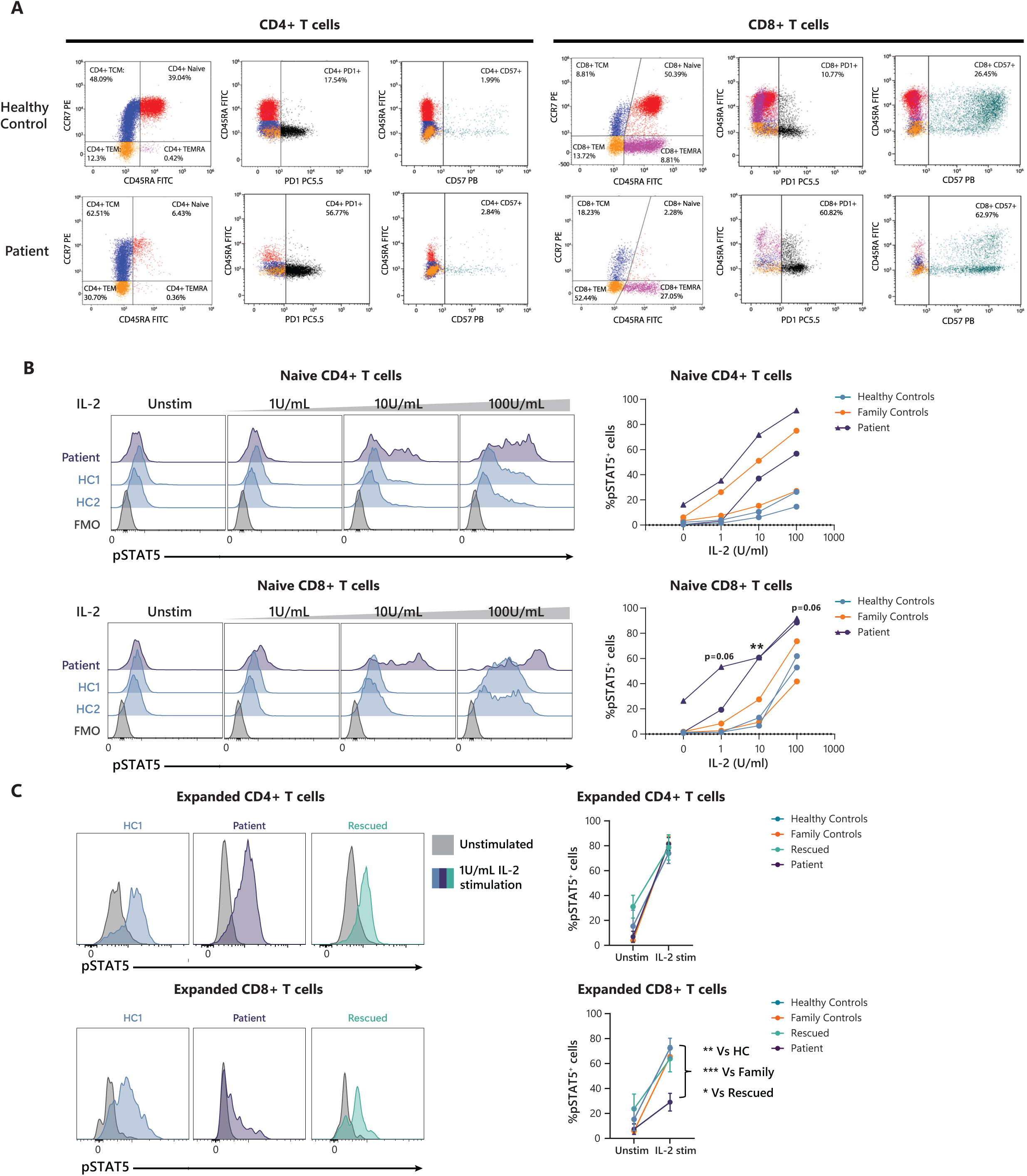
T-cell phenotyping of the index patient. A) T-cell flow cytometry dot plots showing CD4 T and CD8 T-cell-specific comparisons of the patient and one representative age-matched HC. Subset analyses of naïve, TCM, TEM, and TEMRA are shown in the first column, as determined by their expression of CCR7 and CD45RA. Expression levels of the T-cell exhaustion marker PD1^+^ and senescence marker CD57^+^ are presented in columns 2 and 3, respectively, for both CD4 and CD8 T-cells along with the expression of CD45RA. Age-matched reference ranges for quantification of different subsets are presented in Table 1. B) pSTAT5 response in the naïve CD4 T and naïve CD8 T-cell subsets of PBMCs from the patient (purple), family HC (orange), and unrelated HC (light blue) after stimulation with 1, 10, or 100 U/mL IL-2. Flow cytometry histograms of different IL-2 doses are presented on the left. pSTAT5^+^ cells were quantified by gating after unstimulated HCs. Two different time points of patient samples are represented with different symbols: triangles (time point 1) and circles (time point 2). \*\**P* < .01, unpaired *t* test at each dose followed by the Holm– Šídák multiple comparison test. C) pSTAT5 abundance in expanded T-cells from the patient and controls (HC and family) was measured by flow cytometry with gating for CD4 and CD8 T-cell subsets. Abundance was measured before and after stimulation with 1 U/mL IL-2 for 15 minutes (*n* = 3). A histogram from one representative experiment is shown here. pSTAT5^+^ cells were determined by gating at the unstimulated patient sample in each experiment. \**P* < .05, \*\**P* < .01, \*\*\**P* < .001, unpaired *t* test at each dose followed by the Holm– Šídák multiple comparison test. FMO, fluorescence minus one; TCM, central memory T-cell; TEM, effector memory T-cell; TEMRA, effector memory RA^+^ T-cell.

**Table 1:**
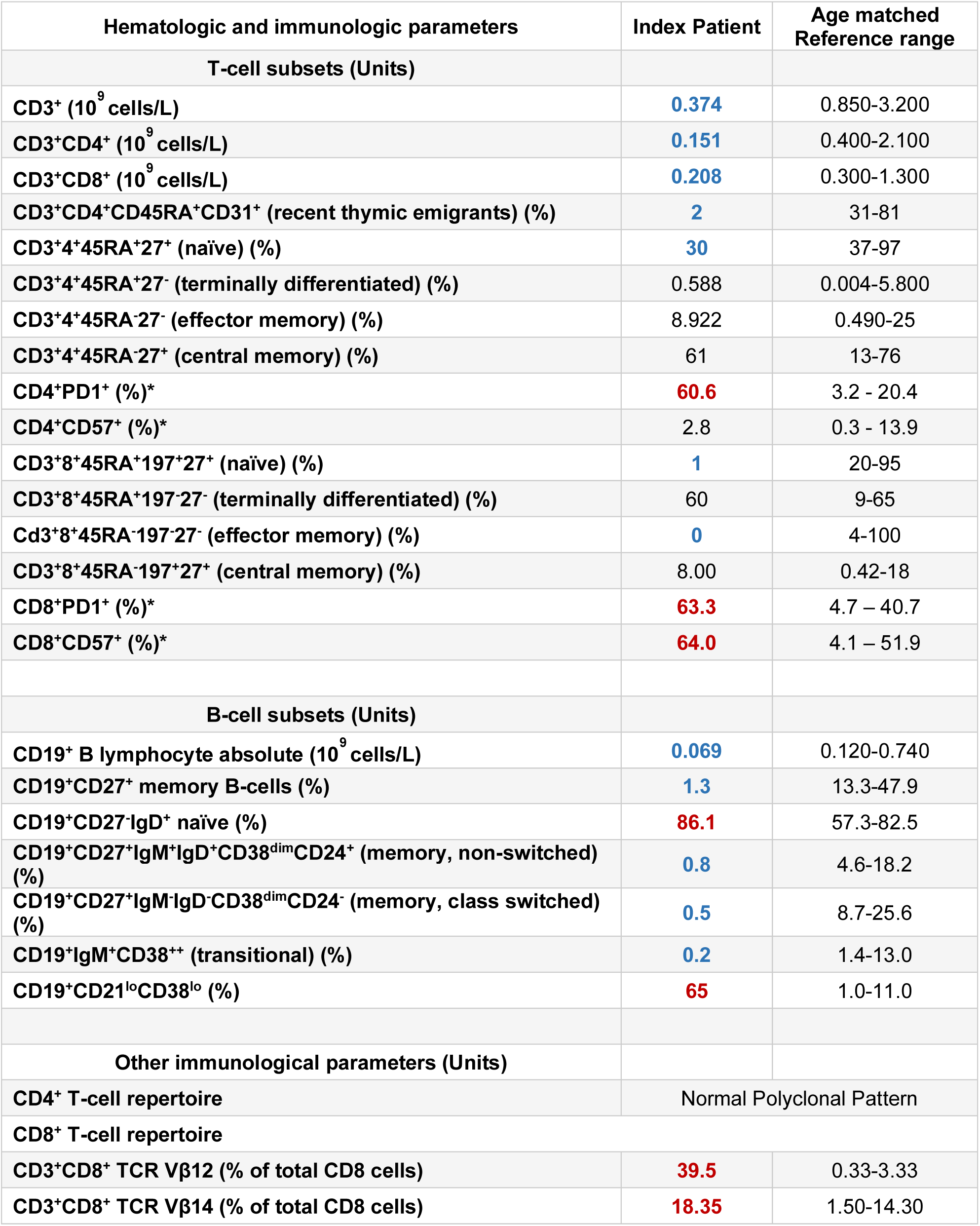
Immunophenotyping of Index patient lymphocytes in early adolescence (10-15 years-of-age). Red denotes high values and blue denotes low values. The * indicates values with a reference range that is based on adult healthy controls.

## DISCUSSION

Here, we reported the clinical, epigenetic, and biochemical findings of the first case of autosomal recessive ASXL1 deficiency in a female with combined immunodeficiency and EBV-associated Hodgkin lymphoma. Single-patient studies account for approximately one-third of newly identified monogenic immune disorders,^1^ requiring methodical validation to establish causality. Following the guidelines proposed by Casanova et al. for studies in single patients,^41^ we showed that the *ASXL1* variants identified in the patient are rare, monogenic, and have complete penetrance. Studies in primary patient cells demonstrated decreased ASXL1 protein levels, dysregulated DNAm, and epigenetic age acceleration. Finally, we showed that epigenetic and immunological defects in the patient’s cells were rescued by transduction of WT *ASXL1*. The patient’s rare and severe clinical presentation provided further support for an underlying genetic cause.^41^ She had a combined immunodeficiency, with T-cell lymphopenia from early childhood followed by progressive loss of B-cells. After receiving live MMR vaccine, she developed vaccine-strain rubella-positive granulomas.

From a hematology/oncology perspective, she had persistent macrocytosis and mild bone marrow hypocellularity, and developed chemorefractory advanced Hodgkin lymphoma. Based on our findings, patients presenting with this unique clinical profile should be evaluated for autosomal recessive ASXL1 deficiency.

Epigenome-wide association studies (EWAS) showed significant alterations of DNAm in patient T-cells, PBMCs, and WB, but not buccal swabs; DNAm decreased predominately in T-cells and WB, but was increased in PBMCs. The contrasting significance and directionality of EWAS results across tissue types suggests cell type-specific impacts of the *ASXL1* variants on global DNAm. This was further supported by the observation of pronounced increase in epigenetic age and reduced telomere length exclusively in lymphocytes. Global hypomethylation and focal hypermethylation are key features of aging, as epigenetic marks are ineffectively maintained through S phase of mitosis.^42^ We showed the patient’s *ASXL1* LoF variants drive epigenetic features with DNAm loss and increased epigenetic age in T-cells. This is supported by recent findings showing altered chromatin accessibility and DNAm inhibition in *ASXL1* KO CD8 T-cells^43^, and high epigenetic age differences in BOS patients with *ASXL1* truncation variants^24^, highlighting the role of *ASXL1* on cellular aging.

Our detailed molecular and clinical evaluation of human ASXL1 deficiency provides further insight into the role of immune dysregulation in cancer predisposition. Global hypomethylation has also been proposed to act as a replicative barrier against highly proliferative and premalignant cells by inducing cellular senescence.^44,45^ In this case, however, T-cell senescence may have played a role in impaired antiviral and antitumor immunity, contributing to the development of EBV-associated Hodgkin lymphoma.^46^ Indeed, recent modeling of an *ASXL1* variant associated with clonal hematopoiesis showed that it promoted the progression of solid tumors via impaired intrathymic T-cell development, naïve-memory T-cell imbalance, and T-cell exhaustion^29^. Loss of *Asxl1* and overexpression of truncated *Asxl1* in mice also lead to limited hematopoietic stem cell self-renewal capacity and decreased peripheral lymphocyte numbers.^22,29,47,48^ Consistent with these findings, we also observed the near-absence of recent thymic emigrants and naïve T-cells, combined with T-cell exhaustion in the patient. The altered epigenetic and immune landscape, advanced cellular aging, CD8 T-cell senescence, and T-cell exhaustion in the context of chronic infection likely all contribute to an oncogenic environment. Some of these mechanisms may also play roles in other hematopoietic malignancies, specifically acute myeloid leukemia, chronic myelomonocytic leukemia, and MDS,^49,50,51^ where somatic *ASXL1* variants are commonly described.^49^ Taken together, *ASXL1* LoF variants may drive the maintenance of a protumor milieu.

The granulomas that emerged in early childhood (0 to 5 years of age) represented a particularly enigmatic feature of this patient’s clinical course. These lesions persisted for more than a decade, were refractory to treatment, and were labeled as idiopathic until recognized as positive for vaccine-strain rubella virus. Rubella virus was first identified in the granulomas of three IEI patients in 2014,^52^ and are now linked to ∼60% of idiopathic cutaneous granulomas in IEIs. Rubella granulomas have been typically found in individuals with ataxia telangiectasia and other related DNA repair disorders.^53,54,55,56^ Our study adds to this relatively new field by suggesting dysregulation of an epigenetic modifier, *ASXL1*, as a cause of vaccine-strain rubella-associated granuloma. The profound T-cell impairments and deficiency described here represent possible underlying mechanisms, as defective T-cell cytotoxicity has been implicated in rubella vaccine-associated granulomas.^56^ Further studies are needed to elucidate the immunological basis of this rare and serious complication.^56^

Heterozygous germline truncating variants of *ASXL1* are known to cause the neurodevelopmental disorder BOS.^57^ The hematologic and immune disorder described here was distinct from BOS in both its molecular and clinical effects. We identified biallelic missense *ASXL1* variants located outside the mutational hotspot associated with BOS. The DNAm signature in our patient also does not overlap with that seen in BOS patients^24^. A recent study further showed no impairment of ASXL1 protein level in BOS patients, contrasting our findings.^23^ Studies in mouse models indicated that *Asxl1*^−/−^ mice show developmental abnormalities and failure to thrive, recapitulating elements of the BOS patient phenotype,^22,47^ while *Asxl1^−/+^* mice develop an MDS-like phenotype possibly attributable to *Asxl1* haploinsufficiency.^22^ Given these findings, BOS-causing *ASXL1* variants likely have different functional impacts than those in our patient, which may contribute to the distinct phenotype observed. Future studies directly comparing samples from patients with autosomal recessive ASXL1 deficiency to those with BOS may further elucidate the mechanistic differences between these two diseases.

This study had many strengths, notably the detailed clinical and immunological description of the first human case of biallelic ASXL1 deficiency and well-controlled epigenetic profiling with family controls and a population reference data set across multiple tissues. However, there were also limitations. First, this was a single-patient study, and ultimately the full phenotype of human ASXL1 deficiency will only be defined through diagnosis of more patients. Second, while we observed global differential DNAm in patient whole blood and PBMCs, we explored the effect of WT *ASXL1* transduction only in patient T-cells. Sample limitations restricted us from working on other cell types (eg, neutrophils) in a controlled setting. Similar to previous findings, we showed the effects of *ASXL1* variants to be highly cell type-specific,^48^ and therefore examining different cellular models could elucidate the roles of ASXL1 in hematopoiesis and disease. Finally, although our patient has benefitted clinically from allogeneic HSCT, follow-up was only 6 months at the time of writing, so the observed benefits should be interpreted with caution.

In conclusion, our study identified a novel human IEI caused by germline biallelic deleterious *ASXL1* variants. These observations will empower diagnosis of future patients with ASXL1 deficiency, and expand the clinical spectrum of disease associated with LoF *ASXL1*. We recommend genetic testing at the first sign of combined immune deficiency as a powerful precision medicine approach to guide diagnosis and inform therapeutic decision-making processes.

## Data Availability

The data produced in the present study is available upon reasonable request to the corresponding author.

## Acknowledgements

We are grateful to our colleagues, Karlie Edwards, Chaini Konwar, Hilary Brewis, Hannah-Ruth Engelbrecht, and Alan Kerr, who critically contributed to the flow and style of the manuscript. Thanks are also due to Nicole Gladish who facilitated the downloading and merging of ChromHMM data from the Roadmap Epigenomics database. We are also grateful for the help in patient management and organization of the investigation effort we received from Anne K Junker.

## Contribution

Contribution described using the CRediT taxonomy.

Contributions=Conceptualization, Methodology, Formal Analysis, Investigation, Visualization, Writing-original draft preparation, Writing-reviewing and editing (Maggie P Fu, Mehul Sharma).

Contributions= Investigation, Resources, Writing-reviewing and editing (Sarah Merrill, Pariya Yousefi, Ryan Tan, Bhavi P Modi, Kate Del Bel, Rebecca J Deyell, Jacob Rozmus, Wingfield Rehmus, Kyla J Hildebrand, Elliot James, Géraldine Blanchard-Rohner, Susan Lin, Kevin E Shopsowitz, Audi Setiadi, Jefferson Terry, Anna F Lee, Britt I Drögemöller, Allison Matthews, Maja Tarailo-Graovac, Laura Sauvé, Hana Mitchell, Julie S Prendiville, Julie L MacIsaac, Kristy Dever, David T S Lin, Mandy Meijer, Colin J D Ross, Simon R M Dobson, Suzanne M Vercauteren, Wyeth W Wasserman, Clara D M van Karnebeek, Margaret L McKinnon).

Contributions=Conceptualization, Methodology, Investigation, Validation, Visualization, Resources, Funding acquisition, Project administration, Supervision, Writing-original draft preparation, Writing-reviewing and editing (Michael S Kobor, Stuart E. Turvey, Catherine M Biggs).

