## Supplementary information for "Human ASXL1 Deficiency Causes Epigenetic Dysfunction, Combined Immunodeficiency and EBV–Associated Hodgkin Lymphoma"

### SUPPLEMENTARY METHODS

#### Sanger sequencing

Briefly, standard sequencing of genomic DNA from all individuals of the family was performed to confirm genotype segregation with the disease in the family using the following primers: Var1: Forward 5′-GAATGACTGCCTTGCTTTAAGA-3′, Reverse 5′-AAAGGTTAGTCCCTAGTGCC-3′; Var2: Forward 5′-TGCCAGATAGCTGTGAAACA-3′, Reverse 5′-CACATTTGGAGATGAGAAACGA-3′.

#### Identification of candidate variant

Based on family history and pedigree, once a candidate variant list was generated by the pipeline, variants identified through the de novo, autosomal recessive, and compound heterozygous inheritance models were first assessed. All high-impact, putative loss-of-function (LoF) variants were reviewed. Next, the candidate variant list was screened for variants in genes associated with inborn errors of immunity (IEI), especially those previously linked with vaccine-strain rubella-associated granulomas. Genes associated with DNA repair deficiency disorders as well as telomeropathies were also investigated.

In addition, a phenotype-driven approach was also employed to identify relevant variants in genes that may be associated with clinical features (using associated MeSH and HPO terms) presented by the proband, such as multiple skin lesions, necrotizing granulmatous inflammation (postvaccine rubella strain identification), idiopathic chronic macrocytosis, low T-cell count, high B-cell count, mild megablastoid changes, mild dysplastic features, infections, pneumonia, failure to thrive, learning disability, and subtle dysmorphic features.

#### Generation and maintenance of primary patient T-cells and fibroblasts

T-cells were treated with exogenous IL-2 (10 U/mL) and ImmunoCult™ Human CD3/CD28 T Cell Activator (Cat # 10971; Stemcell, Vancouver, BC, Canada) for 12 days and maintained at a concentration of $10^6$ cells/mL every 2 days. After 12 days, exogenous IL-2 concentration was increased to 1000 U/mL.

To grow fibroblasts, patient skin was incubated with complete DMEM composed of 10% heat-inactivated FBS (Gibco-BRL Life Technologies, Grand Island, NY, USA; Life Technologies, Gaithersburg, MD, USA), 2mM L-glutamine (HyClone, Thermo Fisher Scientific, Waltham, MA, USA), 1mM sodium pyruvate (Gibco-BRL Life Technologies), and 1× Antibiotic-Antimycotic (Gibco-BRL Life Technologies, Thermo Fisher) for 3 weeks at 37°C or until a confluent monolayer of fibroblasts had formed. Primary fibroblasts were then lifted and frozen at −80°C for future assays.

**Stable expression of ASXL1 using a lentivirus vector**

To generate lentivirus vectors, wild-type *ASXL1* (NM_015338) plasmid (Cat#: RG219354; OriGene Technologies, Rockville, MD, USA) was cloned into a GFP-tagged Lenti vector (Cat#: PS100071; OriGene Technologies) using *Asc*I (Cat#: R0558L) and *Mlu*I-HF (Cat#: R3198L) (both from New England Biolabs, Ipswich, MA, USA). The plasmid was packaged using third-generation packaging plasmids and transfected into HEK293T cells. Culture medium was collected, centrifuged, filtered, concentrated, and stored at −80°C until use. Expanded T-cells were infected with lentiviral particles and expanded in complete RPMI-1640 (GE Healthcare, Chicago, IL, USA) supplemented with 10% FBS for 9 days.

**Intracellular flow cytometry in PBMCs, expanded T-cells, and primary fibroblasts**

Expanded T-cells were first washed and stained with CD4 BUV563 (Clone: SK3, Cat# 612912; BD Biosciences, Franklin Lakes, NJ, USA), CD8 BUV395 (Clone: RPA-T8, Cat# 563796; BD Biosciences), and Fixable Viability Stain 780 (Cat#565388; BD Biosciences) for 15 minutes. Cells were then fixed (fibroblasts were lifted, spun, and fixed) using BD Cytofix (Cat# 554655; BD Biosciences) for 20 minutes at room temperature and permeabilized using Perm III for 30 minutes on ice (Cat# 558050; BD Biosciences). The cells were then stained with either primary antibody for ASXL1 (Cat# MA5-36238; Thermo Fisher Scientific) or mouse IgG1 isotype control for 1 hour at room temperature in 1× Perm/Wash (Cat# 554723; BD Biosciences). Intracellular staining was performed separately for H2AK119ub (Cat# 8240S; Cell Signaling Technology, Danvers, MA, USA) in expanded T-cells. Subsequently, the cells were washed and stained with goat anti-mouse secondary antibody (Alexa Fluor™ 405, Cat#: A-31553; Thermo Fisher Scientific) for 30 minutes at room temperature. ASXL1 expression was then measured using a BD FACSymphony flow cytometer (BD Biosciences) and analyzed using FlowJo software (BD Biosciences).

Detection of pSTAT5 was performed similar to the protocol outlined above in expanded T-cells and in primary patient peripheral blood mononuclear cells (PBMCs). After permeabilization, cells were stained with mouse anti-human pSTAT5 (pY694) PE-Cy7 (Cat# 560117; BD Biosciences). The following antibodies were used for extracellular staining of PBMCs: CD3 FITC (Clone: UCHT1, Cat# 555332; BD Biosciences), CD4 BUV563 (Clone: SK3, Cat# 612912; BD Biosciences), CD8 BUV395 (Clone: RPA-T8, Cat# 563796; BD Biosciences), CD27 BV605 (Clone: L128, Cat#: 562655; BD Biosciences), CD45RA BV421 (Clone: HI100, Cat#: 562885; BD Biosciences).

**Clinical-grade flow cytometry**

Patient clinical immunophenotyping was carried out on whole blood at an accredited clinical flow cytometry laboratory according to standard protocols. Briefly, whole blood was collected and stained using dried antibody reagents (DURAClone tubes; Beckman Coulter, Brea, CA, USA) according to the manufacturer's recommendations. Panels focused on the analysis of T-cell subsets (DURAClone IM T Cell Subsets, Cat#: B53328; Beckman Coulter) and B-cell subsets (DURAClone IM B Cell, Cat#: B53318; Beckman Coulter). Samples used for the B-cell subsets were washed three times prior to staining. Aliquots of 100 μL of fresh blood were added to the dried reagent tubes and incubated in the dark at room temperature for 15 minutes prior to lysis with ImmunoPrep Reagent System (Beckman Coulter). Samples were then fixed with 1.0 mL of 1% paraformaldehyde prior to flow cytometry. All flow data were acquired on a Navios flow cytometer and then analyzed using Kaluza software (Beckman Coulter).

**DNA extraction for DNA methylation array analysis**

Whole blood was collected in Vacutainer® CPT™ Cell Preparation Tubes (Becton Dickinson, Franklin Lakes, NJ, USA). PBMCs were then isolated by standard Ficoll-Plaque (GE Healthcare) density centrifugation. Buccal swabs were collected with Isohelix Buccal Swabs (Cell Projects Ltd., Kent, UK) and Oragene DNA sample collection kits (DNA Genotek, Kanata, ON, Canada), respectively. Genomic DNA was extracted from whole blood, PBMCs, and expanded T-cells with DNeasy kits (Qiagen, Germantown, MD, USA). Buccal DNA was extracted with Isohelix Buccal DNA Isolation kits (Cell Projects Ltd.). Extracted DNA (750 ng) was subjected to bisulfite conversion with an EZ-96 DNA Methylation kit (Zymo Research, Irvine, CA, USA), and aliquots of 160 ng were assayed with Infinium HumanMethylationEPIC BeadChip arrays (Illumina, San Diego, CA, USA).

**Sample selection for pediatric population reference set**

We gathered all cohorts in whole blood, PBMCs, and dried blood spots (DBSs), and excluded cord blood samples. To ensure consistent data preprocessing and minimization of batch effects, we used only cohorts for which raw IDAT files were available in the database. The methylation data and metadata were then compiled manually. Missing metadata were curated from the GEO data set publications and included variables such as age, sex, tissue, ethnicity, and disease status. For disease status, we used the designation of “healthy control” (HC) for samples collected from individuals without persistent severe health conditions, as described in the cohort metadata and the associated publication. Only cohorts that included age information and had samples in the pediatric age range were retained. After quality control (QC), outlier removal, age matching (6–18 years), and selection for HCs, 725 buccal samples, 452 WB samples, 285 PBMC samples remained.

**Epigenetic age analysis**

The *DNAmAge* function in the `methylclock` package was used to estimate all DNAm ages in parallel.[1] Epigenetic age acceleration (EAA) was calculated by fitting regression of epigenetic age (EA) to chronological age (CA) and extracting the residual, representing the component of EA that could not be explained linearly by the CA process, whereas epigenetic age difference (EAD) was defined as the difference between EA and CA.

**Normalization, QC, and imputation of DNA methylation data**

For the analysis of primary tissues, the HC cohorts and patient samples were preprocessed together. All subsequent analyses were performed in R version 4.0.3 (R Foundation for Statistical Computing, Vienna, Austria). The IDAT files were normalized with the *preprocessFunnorm* function in the `minfi` package, which included two types of normalization: Noob (normalized out-of-band) normalization and functional normalization.[2,3] The two normalization steps utilized the readings from control and out-of-band probes to adjust for dye bias from the two-color channels and probe-type bias from Type I and Type II probes on the arrays, respectively. Samples from the 450k and EPIC microarrays were normalized separately, and then combined by subsetting to the shared probes.

QC was performed at both sample and probe levels. Samples with bisulfite conversion rate < 75 or average detection *P*-value > .01 were removed. Three other outlier detection methods—*detectOutlier* from `lumi`,[4] *outlyx* from `wateRmelon`,[5] and Z-test on the first principal component (PC)—were implemented to identify outliers with distinct methylation patterns. Furthermore, sex prediction was performed with the *getSex* function in `minfi`. Samples with mismatches in reported and predicted sex were also removed. Finally, probes in which > 5% of samples had bead count < 3 or an average detection *P*-value > .05 were filtered out.

After QC, the missing values were imputed in a probe-wise manner based on the DNAm patterns of other probes with the lowest Euclidean distance. The method was implemented with the *imputeKNN* function in the `impute` package, where $k = 10$.

**Cell type proportion prediction**

Cell type proportion of whole blood and PBMC DNAm data were estimated from the imputed data with the Houseman reference-based prediction method,[6] as implemented using the *estimateCellCounts2* function from the `FlowSorted.Blood.EPIC` package with the `FlowSorted.BloodExtended.EPIC` reference.[7] In buccal swabs, cell type proportion was predicted with the *hepidish* function in the `EpiDISH` package[8]. We compared the estimated proportion of each cell type between the patient and HCs using the one-sample *t* test.

**Correcting for variances associated with intersample cellular heterogeneity**

Given that the patient shows unique cell type proportions distinct from the HC, it is essential to adjust for cell type-associated DNAm variation to prevent confounding with the disease-associated DNAm signature. In the primary tissues, this was accounted for with a regression model. To overcome the compositional nature of cell type proportion data, we first transformed the proportion into an isometric log ratio followed by principal component analysis (PCA) to extract components that capture the variabilities corresponding to cell type differences, yet are not bounded or correlated. We then fitted a regression model of the DNAm beta value as a function of the cell type PCs[9] and extracted the residuals as the cell type-adjusted DNAm values. For the analysis in T-cells, we accounted for the CD8 proportion directly as a covariate in our EWAS.

**Batch correction**

Given that we only included samples for which raw IDAT files were available, we also had access to the chip ID and position at which each sample was run. We identified three known sources of batch effect, ie, the array platform (450k or EPIC), chip ID, and chip row. Therefore, we adjusted the data for these three known batch effects with the *ComBat* function in the `sva` package,[10] which uses an empirical Bayesian framework to estimate the batch effect on samples across probes and account for them with a regression model. Each cohort can also be considered as a batch, but that is often confounded with different tissues, disease status, and age range, so we did not include cohort as a known batch variable. We confirmed that the batch effect of the three variables were removed with PCA. Platform ID, chip ID, and chip row were originally strongly correlated with PC1 through PC5 on Pearson's correlation test (multiple test adjusted $P < .001$), and most of the effects were removed by *ComBat*. For the T-cell DNAm data, we accounted for batch in the `limma` regression model, as too few batches were present, and we did not detect a strong batch effect within each time point.

**Differential DNAm analysis of expanded T-cells**

The expanded T-cells for the DNAm arrays were collected over 3 days to capture some biological and technical variability. In the linear regression model, we accounted for concomitant variables in our analysis, including sex, age, and technical batch, to specifically isolate the effect of patient *ASXL1* genotype. We identified 276 769 differentially methylated sites (DMs) with a statistical cutoff with $P_{adj} \leq .05$ and effect size (ES) cutoff with delta-beta > 0.1. The same regression model comparing *rescue vs. control* yielded 208,692 DMs passing the statistical and ES thresholds. We then included flow-derived $CD8^+$ proportion with other concomitant variables in the model to account for cellular heterogeneity. With the new model, we no longer observed any DMs passing the statistical threshold of $P_{adj} < .05$, showing that *ASXL1* variants led to an imbalance in $CD4^+$ and $CD8^+$ T-cell proportions, and that ASXL1 has a strong cell population-specific effect on DNAm pattern. We relaxed the statistical threshold to $P_{adj} < .2$ and detected 141,113 DMs. Downstream analysis are performed with this model and detection threshold.

**Enrichment analysis for differential DNA methylation**

Overrepresentation analyses were conducted using the *gometh* function in the `missMethyl` package to identify enriched ChromHMM chromatin states. The ChromHMM states were exported from the Roadmap Epigenomics database and overlapped with the Illumina annotation file for the EPIC arrays.[11] The method accounts for the background probes on the DNAm arrays to minimize coverage biases. The probes were annotated to the corresponding genes and the enriched pathways with multiple test-adjusted $P < .05$ were identified.

#### Supplementary Clinical Data

Presentation and treatment of EBV-associated Hodgkin lymphoma

Between 10 to 15 years of age the patient presented with a three-month history of epistaxis and gum bleeding. Mildly reduced platelets with elevated mean platelet volume, suggestive of peripheral destruction, prompted an abdominal ultrasound which identified hepatosplenomegaly. A chest CT demonstrated diffuse intrathoracic lymphadenopathy and baseline positron emission tomography (PET)-CT revealed $^{18}$F-fluorodeoxyglucose (FDG)-avid disease above and below the diaphragm, pulmonary nodules, hepatic and splenic lesions with innumerable marrow deposits. An axillary lymph node biopsy confirmed the diagnosis of stage IVA classical Hodgkin lymphoma, mixed cellularity subtype with prominent granulomatous inflammation (**Supp Fig 1D**). She had an associated EBV viremia (EBV viral load 15,529 copies/ml) and immunohistochemistry of Reed-Sternberg cells from her diagnostic lymph node biopsy were positive for EBV-encoded RNA (EBER). Bone marrow aspirate and biopsy showed numerous lymphohistiocytic aggregates, in addition to mild megakaryocytic atypia and mild red blood cell macrocytosis.

She received 2 cycles of ABVD (doxorubicin [adriamycin], bleomycin, vinblastine and dacarbazine) chemotherapy with a partial metabolic response (Deauville 4), and was then switched to AVD with brentuximab-vedotin, the anti-CD30 antibody, for cycles 3 and 4 due to significant myelosuppression and slow early response. Repeat PET-CT done after 4 cycles of chemotherapy showed progressive disease with 2 new FDG-avid lesions (spleen and right humerus). The splenic lesion was biopsied and histology remained in keeping with classical Hodgkin lymphoma. Following tumour board review, she initiated immunotherapy with nivolumab, the anti-programmed cell death protein 1 (PD1) antibody. This was complicated by the development of autoimmune thyroiditis, which was treated with thyroxine. She completed 4 cycles of nivolumab after which PET-CT re-evaluation was indeterminate with persistent FDG-avid lesions potentially related to her chronic skin lesions and an intercurrent axillary seroma, but no definitive disease progression. Six weeks following discontinuation of nivolumab, she began conditioning for a 9/10 matched unrelated donor hematopoietic stem cell transplant with cyclophosphamide, fludarabine and 200cGy total body irradiation. She received mycophenolate mofetil and tacrolimus prophylaxis for graft versus host disease (GVHD) and developed grade 1 GVHD of upper gut and skin which was treated with corticosteroids. She was discharged on day +34 with full donor chimerism, and remains well now 6 months post-transplant with healing of her chronic skin lesions.

#### Supplementary Figure Legends:

**Supp. Figure 1:** A) White blood cell count assessment over time showed normal white blood cell count. B) Immunoglobulin assessment over time showed low IgG until initiation of intravenous immunoglobulin (IVIG), low/normal IgM levels and low/normal IgA levels. C) Skin biopsy performed in childhood (0-10 years of age) from the left thigh. Tissue section stained with hematoxylin and eosin showing subcutaneous necrotizing (*) and nonnecrotizing granulomas (arrowhead). Specific staining for organisms was negative (not shown). Scale bar: 200 μm. D) Micrograph of lymph node indicating juxtaposed granulomatous inflammation (*) and classical Hodgkin lymphoma with Reed–Sternberg cells (arrowheads). Scale bar: 200 μm.

**Supp. Fig. 2:** A) Predicted CADD score (v1.6) and allelic frequency of ASXL1 variants reported in the gnomAD database (v3). All missense (gray), stop-gained (navy blue), or frameshift (magenta) variants are plotted. Patient variants are indicated with a triangle.

**Supp. Fig. 3:** A) Volcano plot of differential DNAm pattern detected in buccal swabs with linear regression. Delta beta represented the effect size (regression coefficient) of the patient status variable after accounting for sample age and sex. Magenta points indicate DMs with increased DNAm in the patient, and turquoise points represent those with decreased DNAm. The vertical gray line represents the delta-beta = 0.05, and the horizontal line corresponds to $P_{adj}$ = 0.05. B) Overlap of differentially methylated CpGs identified in our whole blood analysis and those associated with Bohring–Opitz syndrome in a previous study[13]. C) CA and predicted PedBE EA in the patient and HCs. HC samples are shown in light blue, while patient samples are shown in red. The black line indicates the best fit line using linear regression of all the sample points. The gray area corresponds to the 95% confidence interval. D) Telomere length of patient lymphocytes and granulocytes compared with the reference range in the HC population.

**Supp. Fig. 4:** A,B) Epigenetic age analysis in expanded T-cells, (*n* = 3 for patient and rescued samples). A) Difference in Horvath skin and blood epigenetic age and chronological age. B) Difference in Hannum epigenetic age and chronological age. 0.05, **$P < .01$, ***$P < .001$, one-way analysis of variance with Dunnett's multiple comparison test.

**Supp. Fig. 5: Extended T-cell phenotyping of the index patient.** pSTAT5 response in the A) memory CD4 T and B) memory CD8 T-cell subsets of patient and control PBMCs after stimulation with 1, 10, or 100 U/mL IL-2. Patient (purple), family controls (orange), HCs (light blue). Two different time points of patient samples are presented with different symbols (triangles and circles). C) Oxidative stress was detected in expanded $CD4^+$ and $CD8^+$ T-cells of the patient and controls (family and HC) by flow cytometry using CellROX Deep Red reagent. *$P < .05$, **$P < .01$, one-way analysis of variance and Tukey's post hoc test.
